# Early-Horizon Multimodal ICU Mortality Prediction Without Retraining

**DOI:** 10.64898/2026.05.18.26353392

**Authors:** Alexander Bakumenko, D. Hudson Smith, Janine Hoelscher

## Abstract

Earlier ICU mortality prediction is more clinically useful because it can identify high-risk patients while treatment decisions can still change. Yet most models are trained on data from a fixed time window, so it is unclear whether a model trained on the first 48 hours of ICU data remains reliable when used earlier in the ICU stay. We evaluated a multimodal ICU mortality model trained once at 48 hours and then applied unchanged at 6, 12, 24, and 48 hours on MIMIC-III. The model combines an LSTM for physiological time-series data, a finetuned ClinicalModernBERT model for clinical notes, and a logistic regression fusion layer. Performance remained strong at earlier time points, suggesting that useful mortality prediction is possible earlier in the ICU stay even without retraining. At 6 hours, the model achieved AUROC 0.777 and remained well-calibrated (expected calibration error, ECE 0.038) without any recalibration, and it outperformed both single-modality models at every horizon. The benefit of combining both modalities was most evident at earlier horizons, when physiological data were sparse: agreement between the two specialists dropped by more than half from 48 to 6 hours, while the median contribution from clinical notes increased from 37% to 49%. A Bayesian version of the fusion layer showed that uncertainty decreased for survivors as more data accumulated but remained high for non-survivors; the most uncertain cases were up to 4.9 times more likely to be non-surviving patients. Continuous hourly analyses further showed that clinical notes provide stable context between documentation events. Simply carrying forward the most recent note matched or outperformed note-decay and documentation-gap alternatives. These results suggest that a multimodal ICU mortality model trained on 48 hours of data can provide trustworthy earlier predictions without retraining, while also identifying the cases that remain hardest to interpret.

## 1. Introduction

Early ICU mortality prediction is most useful when intervention is still possible. Machine learning ICU mortality predictors are overwhelmingly trained and evaluated at a single fixed horizon, typically 24 or 48 hours, yet the window where intervention is most actionable is earlier Harutyunyan et al. (2019); Topol (2019). When such a model is applied before its training horizon without adaptation, its trustworthiness, here meaning reliable behavior with calibrated discrimination and quantified per-case uncertainty, cannot be assumed Subbaswamy and Saria (2020). For multimodal models that combine structured physiological time-series with clinical text, the mismatch is even greater: a model trained at 48 h encounters systematically different input distributions at, for instance 6 or 12 h, such as shorter physiological trajectories and fewer available clinical notes, both of which alter the evidence presented to the model Khadanga et al. (2019). This mismatch is not uniform: clinical notes arrive sparsely but often capture broader, longer-lived context, whereas physiological time-series update frequently and primarily reflect current state Zhang et al. (2022). Whether such a model remains trustworthy when applied earlier than its training horizon has not been systematically characterized.

A clinical prediction cannot be trusted on discrimination alone; three properties matter jointly: *discrimination*, so that higher-risk patients can be ranked above lower-risk patients; *calibration*, so that predicted probabilities reflect observed outcome frequencies; and *per-case uncertainty*, so that clinicians can distinguish stable predictions from those that warrant additional scrutiny Combi et al. (2022); Tonekaboni et al. (2019). These properties need not degrade in parallel as horizon shortens. A model may retain rank-order performance while losing calibration, or remain accurate in aggregate while being unreliable for a clinically important subset of episodes. This distinction matters concretely: calibration errors directly distort decision thresholds and risk communication Van Calster et al. (2019), and principled per-case uncertainty has been identified as an essential requirement for machine-assisted medical decision making Begoli et al. (2019). Neither property is well-documented in the multimodal ICU mortality literature. A recent scoping review of multimodal ICU in-hospital mortality studies found that only one-third reported any calibration metric, and trustworthiness analysis across different time horizons was essentially absent Bakumenko et al. (2026). From a pre-clinical standpoint, this is a substantive gap: predictable behavior, calibration, and principled uncertainty quantification have been identified as required properties for any predictive system before clinical evaluation is contemplated Bakumenko et al. (2025b).

We address this gap through a *zero-shot horizon transfer* evaluation paradigm. A multimodal ensemble whose 48-hour behavior has already been established Bakumenko et al. (2025a) is evaluated at 6, 12, 24, and 48 hours without retraining any specialist model and without re-estimating the fusion rule combining the specialist model outcomes. We characterize early-horizon trustworthiness along four axes: (1) discrimination and calibration across horizons; (2) changes in modality complementarity and per-case contribution structure under frozen late fusion; (3) per-case epistemic uncertainty (uncertainty in the model’s own fusion weights) via a Bayesian No-U-Turn Sampler (NUTS) extension of the logistic regression (LR) stacker Hoffman et al. (2014); and (4) continuous hourly sensitivity of predictions to the note imputation strategy under irregular note updates. This paper makes four contributions:

i. We provide a framework for systematically evaluating performance across earlier prediction horizons (6–48 h) without retraining, and show that a frozen multimodal ensemble retains discrimination and calibration while consistently outperforming both unimodal specialists.
ii. We show that modality complementarity is highest at the earliest horizons: clinical notes account for a larger relative share of the fused prediction, while the two specialist outputs are largely uncorrelated. Together, these trends provide a model-grounded explanation for the preserved fusion advantage when physiological evidence is most limited.
iii. We introduce a Bayesian NUTS extension of the LR meta-stacker that preserves point-estimate discrimination while adding per-case epistemic uncertainty, revealing a class-level asymmetry in which survivors stabilize but non-survivors remain persistently uncertain across horizons.
iv. We show that clinical notes act as stable background information: explicitly modeling how notes age over time did not improve predictions, suggesting that notes provide steady contextual signal that remains relevant over time.

### Generalizable Insights about Machine Learning in the Context of Healthcare

Our work offers several insights relevant to multimodal clinical prediction beyond the ICU mortality setting. First, we share a methodology for zero-shot trustworthiness profiling: a frozen model can be characterized across earlier horizons without retraining, identifying where performance, calibration, and per-case reliability remain acceptable before external validation. Second, modalities can be most complementary when information is sparse early on: as the observation window extends and physiological evidence accumulates, the two modalities become progressively more redundant. Multimodal systems should therefore be evaluated across plausible deployment horizons, not only at the training horizon. Third, transparent linear fusion preserves an important interpretability property: a logistic metalearner yields algebraically exact per-case modality attribution, avoiding reliance on post-hoc approximation methods for fusion-level interpretation. Finally, clinical notes can encode stable patient context throughout the ICU stay. Rather than assuming notes become stale over time, this should be evaluated empirically: notes may carry general admission-level information that remains relevant well beyond when they were written.

## 2. Methods

### 2.1. Data, Cohort Construction, and Prediction Task

We use the MIMIC-III clinical database Johnson et al. (2016). Structured physiological time-series are prepared following the benchmark pipeline of Harutyunyan et al. (2019): 17 clinical variables are resampled into hourly bins, missing values imputed via last-observation-carried-forward (LOCF) with per-variable normal-value fallback, and each hour is represented as a 76-dimensional feature vector. Clinical notes are preprocessed following Khadanga et al. (2019): notes without a valid chart time are excluded, and notes are concatenated chronologically. Full architectural and training details for the 48-hour baseline are reported in Bakumenko et al. (2025a). We describe only the horizon-extension protocol here.

The prediction task is binary in-hospital mortality (IHM) fixed to post-48 h through end of stay, evaluated independently using data available at *h* ∈ {6, 12, 24, 48} hours from ICU admission. For a given episode at horizon *h*, the time-series input is formed by retaining only the first *h* hourly vectors and zero-padding the remaining 48 − *h* positions (end-padding). The notes input is the concatenation of all notes charted within *h* hours; no notes from beyond the horizon are used. For each horizon, an ICU stay is included if and only if (i) at least *h* hours of structured measurements are available and (ii) at least one clinical note is charted within the first *h* hours. The train/validation/test partition follows the benchmark split of Harutyunyan et al. (2019); each split is then filtered independently by the horizon-specific criteria. Cohort sizes and IHM prevalence across horizons are summarized in Table 1.

**Table 1:** Horizon-specific cohort sizes and in-hospital mortality prevalence. Prevalence is stable (13.01%–12.73%), indicating no horizon-dependent outcome bias.

| Horizon | $N$ | Positive ($y=1$) | Prevalence |
| --- | --- | --- | --- |
| 6 h | 11,881 | 1,546 | 13.01% |
| 12 h | 14,888 | 1,917 | 12.88% |
| 24 h | 15,554 | 1,995 | 12.83% |
| 48 h | 15,861 | 2,019 | 12.73% |

note is charted within the first $h$ hours. The train/validation/test partition follows the benchmark split of Harutyunyan et al. (2019); each split is then filtered independently by the horizon-specific criteria. Cohort sizes and IHM prevalence across horizons are summarized in Table 1.

The 6-hour cohort (*N* =11,881; Table 1) is the most restrictive, excluding stays shorter than 6 hours and stays without an early note. This cohort serves a dual role: as the discrete 6h evaluation split, and as the fixed cohort for all continuous hourly analyses (Section 2.7).

### 2.2. Multimodal Architecture

The system comprises two independently trained specialist models and a lightweight meta-learner. The *time-series specialist* is a stacked bidirectional LSTM Schuster and Paliwal (1997) (8 units) followed by a unidirectional LSTM Hochreiter and Schmidhuber (1997) (16 units), dropout, and a dense sigmoid output. It processes a 48×76 matrix of 17 clinical variables sampled hourly, each hour represented as a 76-dimensional feature vector following Harutyunyan et al. (2019). A Masking(mask value=0) layer is included in the 48 h base architecture; its role during early-horizon evaluation is described in Section 2.3. The *clinical notes specialist* is ClinicalModernBERT Lee et al. (2025) finetuned end-to-end on in-hospital mortality labels, with an 8192-token context window and a linear classification head on the [CLS] token. It processes all notes charted within *h* hours, concatenated chronologically. Both specialists output a scalar mortality probability *p* ∈ (0, 1) and were trained once on 48-hour MIMIC-III data.

Late fusion is performed by a meta-learner that combines the two specialist outputs (Section 2.4). All model parameters are frozen at evaluation time: the specialists generate predictions from truncated inputs, and the meta-learner combines them using the same parameters learned at 48 h. No component is adapted to any earlier horizon. This is the design constraint that makes zero-shot horizon transfer a meaningful evaluation paradigm.

### 2.3. Zero-Shot Horizon Transfer

Evaluating the ensemble at horizon *h <* 48 h is not a simple input truncation: the LSTM was trained exclusively on 48-timestep sequences, so shorter inputs alter the temporal context it was trained to read; and the clinical notes input must be restricted to notes charted within *h* hours, so no future documentation leaks in.

#### Time-series truncation

For horizon *h*, the time-series input is formed by retaining the first *h* hourly vectors and zero-padding the remaining 48 − *h* positions at the end (end-padding). The `Masking` layer then causes the LSTM to skip zero-padded timesteps entirely; they are excluded from recurrent computation rather than processed as zero-valued inputs, preventing contamination of the hidden state with non-physiological signal. A consequence is that predictions are generated from the hidden state at position *h*−1 rather than position 47: the model operates on only the observed physiology, but from an earlier position in the sequence than it encountered during training. End-padding preserves the temporal alignment learned during training; front-padding breaks it.

#### Clinical notes truncation

The notes input at horizon *h* is the concatenation of all notes charted within the first *h* hours. No notes from beyond the horizon are used. For the discrete evaluation horizons (6, 12, 24, 48 h), notes are extracted at the corresponding cutoff. For the continuous hourly evaluation, the most recent available note is carried forward (Section 2.7).

#### Meta-learner

Fusion weights are fixed at their 48h-trained values. No horizon-specific re-estimation is performed at any stage. Horizon evaluation thus tests both the specialists’ ability to predict from incomplete evidence and the meta-learner’s robustness to the positional shift in temporal context.

### 2.4. Stacking Algorithm Selection

We evaluated the generalization of six meta-learner algorithms on the held-out test set: logistic regression (LOGREG), uniform averaging (AVG), gradient boosted trees (GBM), random forests (RF), multilayer perceptron (MLP), and XGBoost (XGB) Wolpert (1992); Breiman (1996); Bakumenko et al. (2025a). Each candidate used the same two standardized specialist logits, evaluated independently at all four horizons. LOGREG and AVG were the top tier at every horizon, while GBM, RF, MLP, and XGB showed consistently lower AUPRC (Appendix Figure 6); with only two meta-features, higher-capacity stackers fail to improve discrimination.

While LOGREG and AVG showed similar discrimination across horizons, we focus on LOGREG for two architectural advantages: (i) exact additive per-case modality decomposition (Section 2.6); and (ii) direct Bayesian extension via NUTS for per-case epistemic uncertainty without changing point predictions (Section 2.5).

### 2.5. NUTS Meta-Stacker and Per-Case Epistemic Uncertainty

#### Transition from logistic regression to Bayesian No-U-Turn Sampler

The logistic meta-learner described in Section 2.4 operates deterministically: given fixed weights *w*^*^ learned at 48 h, it maps specialist logits to a single mortality probability per episode. However, it provides no information about how confident that prediction is for a given episode. To characterize fusion-level epistemic uncertainty without altering the primary model, we replace the point-estimate *w*^*^ with a posterior distribution *p*(*w* | 𝒟), which propagates weight uncertainty to a per-case predictive interval. All other design choices (specialist architecture, fusion in logit space, fixed 48h training) are unchanged. All performance results in Sections 3.1 and 3.2 are based on the deterministic LR stacker.

#### Base specialists and logit-space meta-features

We extend the ensemble from Sections 2.2–2.3 with a fusion-level uncertainty component, keeping all specialist parameters fixed. At each prediction horizon *h* ∈ {6, 12, 24, 48} h, each specialist outputs a probability *p*_TS,*h*_ ∈ (0, 1) and *p*_CN,*h*_ ∈ (0, 1) for in-hospital mortality. To preserve the interpretable additive structure of the stacker, we operate in logit space. Let logit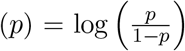. We define the 2D meta-feature vector *x*_*h*_ = [logit(*p*_TS,*h*_), logit(*p*_CN,*h*_)]^T^ and standardize it to *z*_*h*_ using the empirical mean *µ* and element-wise standard deviation *s* computed from the 48-hour training meta-features.

#### Bayesian logistic meta-learner trained once at 48h

We replace the deterministic meta-learner with a Bayesian logistic regression. For the 48-hour training episodes 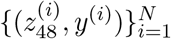 we place weakly informative Gaussian priors on the modality weights *w ∼*𝒩 (0, 1.5^2^*I*) and intercept *b* ∼ 𝒩 (0, 1.0^2^), following standard weak-prior practice for lo-gistic regression Gelman et al. (2008). The likelihood for the binary mortality outcome *y*^(*i*)^ ∈ {0, 1} is:

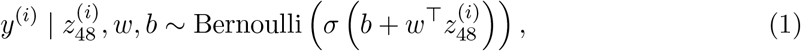

where *σ*(·) is the sigmoid function. Posterior inference is performed with the No-U-Turn Sampler (NUTS), yielding *S* = 1000 posterior samples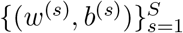.

The posterior is learned *once at 48h* and then *reused across horizons*. Horizon trends in uncertainty therefore reflect changes in input evidence (specialist logits under truncation), not horizon-specific re-estimation of fusion weights. This preserves the zero-shot transfer property of the full system.

#### Posterior predictive inference

For a test episode at horizon *h* with standardized logits *z*_*h*_, we compute 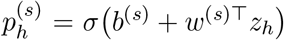for *s* = 1, …, *S*, and use the posterior predictive mean 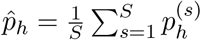 as the point estimate.

#### Epistemic uncertainty metric

We quantify epistemic uncertainty per episode as the width of a central posterior predictive interval on the probability scale:

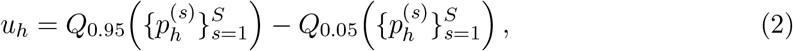

where *Q*_*q*_(·) denotes the empirical *q*-quantile across posterior samples. The distribution of *u*_*h*_ is reported by horizon overall and stratified by true label *y* ∈ {0, 1} using quantiles and violin plots.

Note that two interval types appear throughout: bootstrap 95% CIs on AUROC and AUPRC (*B*=1000 resamples) quantify evaluation-sample variance, reflecting how metrics would vary across test sets drawn from the same population. Posterior predictive intervals *u*_*h*_ (Eq. 2) are distinct; they quantify parameter-level uncertainty per episode, characterizing how much the fused probability for a given patient varies across the posterior over fusion weights.

#### Performance evaluation

For each horizon, we evaluate AUROC and AUPRC on the test set using 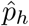, with 95% confidence intervals via nonparametric bootstrap (*B* = 1000 resamples) Tibshirani and Efron (1993).

### 2.6. Modality Attribution Decomposition

The LR meta-learner’s additive linear structure enables an exact, per-case decomposition of the fused prediction into modality-specific contributions: not a post-hoc approximation, but an algebraic identity (Eq. 5 in Section 3.2). The effective raw-logit weights *w*_TS_ and *w*_CN_ are obtained by dividing the standardized-space coefficients *β* by the corresponding training standard deviations *s*; the fused log-odds then decompose exactly as *b*_eff_ + *c*_TS_ + *c*_CN_, where 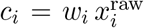 is the signed contribution of modality *i* in raw log-odds units, and *x*_TS_, *x*_CN_ denote the un-standardized specialist logits. Positive values push the prediction toward higher mortality odds; negative values push toward lower.

Two derived metrics characterize fusion behavior across horizons. *Signed contributions c*_TS_ and *c*_CN_ capture the direction and patient-level heterogeneity of each modality’s influence. *Unsigned share* |*c*_CN_|*/*(|*c*_TS_| + |*c*_CN_|), summarized as a per-episode median, captures relative modality dominance. Specialist logit correlation *r*(*x*_TS_, *x*_CN_), computed on raw specialist logits rather than contributions, quantifies information redundancy between specialists independently of fusion reweighting. Together, these three quantities provide the evidence base for the modality complementarity analysis (Section 3.2).

### 2.7. Continuous Hourly Evaluation Under Documentation Sparsity

We hypothesize that clinical notes may encode stable admission-level context, including presenting diagnosis, initial clinician assessment, and chronic conditions, that remains predictive between documentation events rather than behaving as a rapidly decaying signal. To test this, we evaluated the frozen multimodal ensemble hourly from hour 6 through hour 48 on the fixed 6h cohort (*N* =11,881), holding cohort composition constant to isolate evidence accumulation from population shifts.

#### Method 1: Last-Observation-Carried-Forward (LOCF, null model)

Notes are cumulative documents: the 12h note contains all text from the first 12 hours, not only hour 12. We exploit this structure directly: the 6h note is used for hours 6–11, the 12h note for hours 12–23, the 24h note for hours 24–47, and the 48h note for hour 48. LOCF is the null model: if notes persist as stable contextual inputs, this simple strategy should suffice.

#### Method 2: Exponential decay (post-hoc rescaling, no retraining)

To test whether note contributions should diminish monotonically as documentation age grows, we modulate the CN logit at inference time:

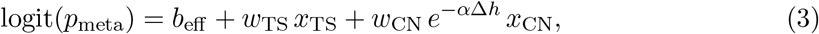

where Δ*h* is hours elapsed since the most recent note and *α* = ln 2*/t*_1_*/*_2_ is the decay rate corresponding to half-life *t*_1*/*2_. We test three half-lives (*t*_1*/*2_ ∈ {6, 12, 24} h). This is post-hoc rescaling only; no meta-learner parameters are re-estimated.

#### Method 3: Additive staleness feature (stacker retrained)

Documentation timing could carry independent predictive signal, as a proxy for monitoring intensity or care-escalation patterns, even if note content itself does not decay. We test this by adding Δ*h* as a third meta-feature and retraining the stacker on validation predictions:

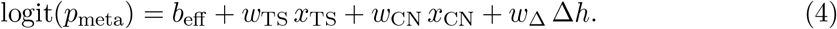

The learned coefficient *w*_Δ_ directly quantifies whether documentation gaps carry predictive signal independent of note content. Results are reported in Section 3.4.

## 3. Results

### 3.1. Discrimination and Calibration Are Preserved Without Retraining

#### Stacking algorithm selection

A comparison of six stacking algorithms (Section 2.4) confirms that LOGREG and AVG dominate at every horizon (Appendix Figure 6); LOGREG is selected over AVG for its exact per-case modality decomposition. LOGREG’s two-weight-plus-intercept structure also motivates the Bayesian extension in Section 3.3: placing a posterior over this parsimonious model adds per-case epistemic uncertainty without changing the predictive structure.

#### Discrimination

We evaluated whether a logistic-regression stacker trained on 48-hour multimodal data retains meaningful discrimination at earlier horizons under a strict zero-adaptation constraint: no retraining, no recalibration, and no horizon-specific tuning. The ensemble achieves AUROC 0.891 and AUPRC 0.565 at 48h, and AUROC 0.777 and AUPRC 0.306 at 6h (Table 2), outperforming both specialists at every horizon by an average of +0.018 AUROC and +0.033 AUPRC. Figure 1 shows AUPRC and ECE confidence intervals across horizons. Performance degrades smoothly (AUROC drop 48h→6h: ensemble −0.114, LSTM −0.162, ft-CMB −0.117), with LSTM losing more AUROC and ft-CMB losing more AUPRC (LSTM: −0.226; ft-CMB: −0.255), an asymmetric degradation examined mechanistically in Section 3.2.

**Table 2:** Discrimination and calibration performance by time horizon for the multimodal ensemble and both modality specialists. All metrics on the held-out test set with 95% bootstrap CIs (*B* = 1000). ECE Pre/Post: expected calibration error before/after recalibration.

| Model | Horiz. | AUROC | AUPRC | Brier (Pre) | Brier (Post) | ECE (Pre) | ECE (Post) |
| --- | --- | --- | --- | --- | --- | --- | --- |
| Ensemble | 48h | 0.891 [0.872,0.909] | 0.565 [0.503,0.625] | 0.068 | 0.068 | 0.022 | 0.022 |
|  | 24h | 0.853 [0.831,0.875] | 0.481 [0.420,0.538] | 0.076 | 0.076 | 0.026 | 0.026 |
|  | 12h | 0.805 [0.778,0.831] | 0.382 [0.318,0.447] | 0.084 | 0.084 | 0.034 | 0.034 |
|  | 6h | 0.777 [0.741,0.809] | 0.306 [0.243,0.367] | 0.084 | 0.084 | 0.038 | 0.038 |
| TS<br>(LSTM) | 48h | 0.855 [0.832,0.877] | 0.485 [0.425,0.545] | 0.075 | 0.075 | 0.029 | 0.024 |
|  | 24h | 0.807 [0.778,0.832] | 0.380 [0.319,0.441] | 0.086 | 0.083 | 0.048 | 0.030 |
|  | 12h | 0.751 [0.719,0.782] | 0.319 [0.261,0.376] | 0.097 | 0.088 | 0.073 | 0.027 |
|  | 6h | 0.693 [0.654,0.732] | 0.259 [0.201,0.320] | 0.104 | 0.089 | 0.101 | 0.035 |
| CN<br>(ft-CMB) | 48h | 0.876 [0.856,0.895] | 0.526 [0.462,0.586] | 0.105 | 0.072 | 0.133 | 0.024 |
|  | 24h | 0.839 [0.816,0.862] | 0.460 [0.402,0.519] | 0.110 | 0.078 | 0.126 | 0.027 |
|  | 12h | 0.782 [0.753,0.810] | 0.344 [0.287,0.403] | 0.114 | 0.087 | 0.105 | 0.035 |
|  | 6h | 0.759 [0.724,0.790] | 0.271 [0.213,0.327] | 0.105 | 0.087 | 0.083 | 0.037 |

**Figure 1:**
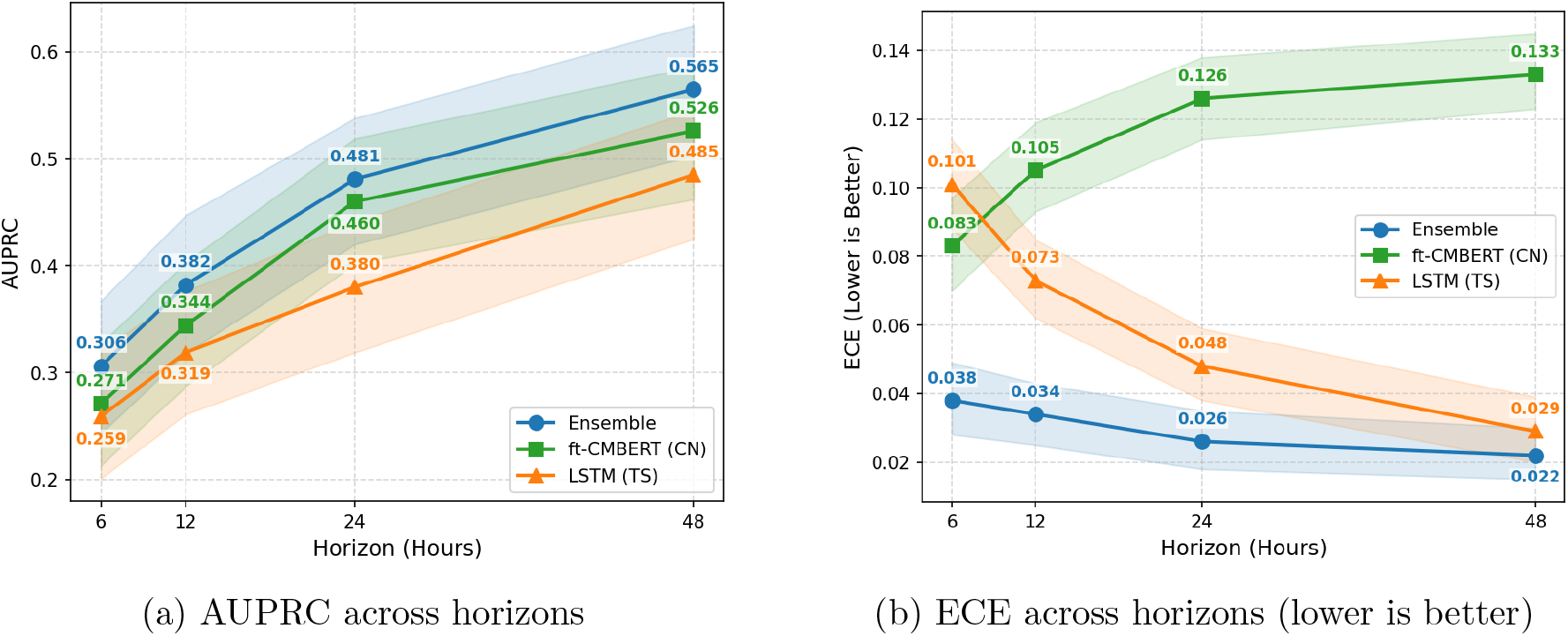
AUPRC (left) and ECE (right) across horizons for the multimodal ensemble and both specialists. Shaded bands show 95% bootstrap confidence intervals (*B* = 1000). Left: the ensemble (blue, circle) outperforms ft-CMB (CN, green) and LSTM (TS, orange) at every horizon; ft-CMB degrades faster than LSTM in AUPRC (48h→6h: ft-CMB −0.255, LSTM −0.226). Right: the LR ensemble maintains ECE ≤ 0.038 with identical pre- and post-calibration values; both specialists require recalibration at early horizons. AUROC and Brier values by horizon are in Table 2; precision–recall and ROC curves are in Appendix Figure 5.

#### Calibration

The LR ensemble requires no post-hoc recalibration at any horizon: ECE is 0.038 at 6h, 0.034 at 12h, 0.026 at 24h, and 0.022 at 48h, with pre- and post-calibration values identical throughout (Table 2). Both specialists require post-hoc recalibration. LSTM is most miscalibrated at early horizons (pre-calibration ECE 0.101 at 6h, 2.7× the ensemble at the same horizon); ft-CMB shows the opposite pattern (ECE 0.133 at 48h), reflecting systematic overconfidence that recalibration corrects substantially. Brier scores follow the same ordering (Table 2). These results indicate that ensemble outputs are directly interpretable as mortality probabilities across 6–48h without additional recalibration. The smallest early-horizon agreement-defined subgroup (6h Agree High, *n* = 33) showed unstable subgroup-level calibration, consistent with limited sample size and distribution shift in rare high-risk consensus cases (see Appendix Table 7).

### 3.2. Modality Complementarity Increases at Shorter Horizons

Table 2 shows that the LSTM specialist performs substantially worse than the notes specialist at earlier horizons: AUROC falls from 0.855 to 0.693 for TS versus 0.876 to 0.759 for CN, yet the LR ensemble consistently exceeds both. This asymmetric degradation cannot be explained by either specialist alone; we therefore examine the fusion dynamics.

We hypothesized that modality complementarity, the degree to which specialists capture distinct, non-redundant information, would increase at shorter horizons, providing a model-grounded account of the preserved fusion advantage when physiological evidence is most limited. The unsigned CN share, |*c*_CN_|*/*(|*c*_TS_| + |*c*_CN_|) per episode, increases monotonically as horizon shortens: median share is 0.370 at 48h, 0.394 at 24h, 0.438 at 12h, and 0.486 at 6h (Figure 2). At 6h, clinical notes and physiological signals contribute approximately equally to the fused prediction. This shift is a structural consequence of the late-fusion architecture under fixed meta-learner weights, not horizon-specific reweighting: as horizon shortens, the LSTM’s output logit compresses toward zero while LR weights remain fixed, so CN gains relative influence.

**Figure 2:**
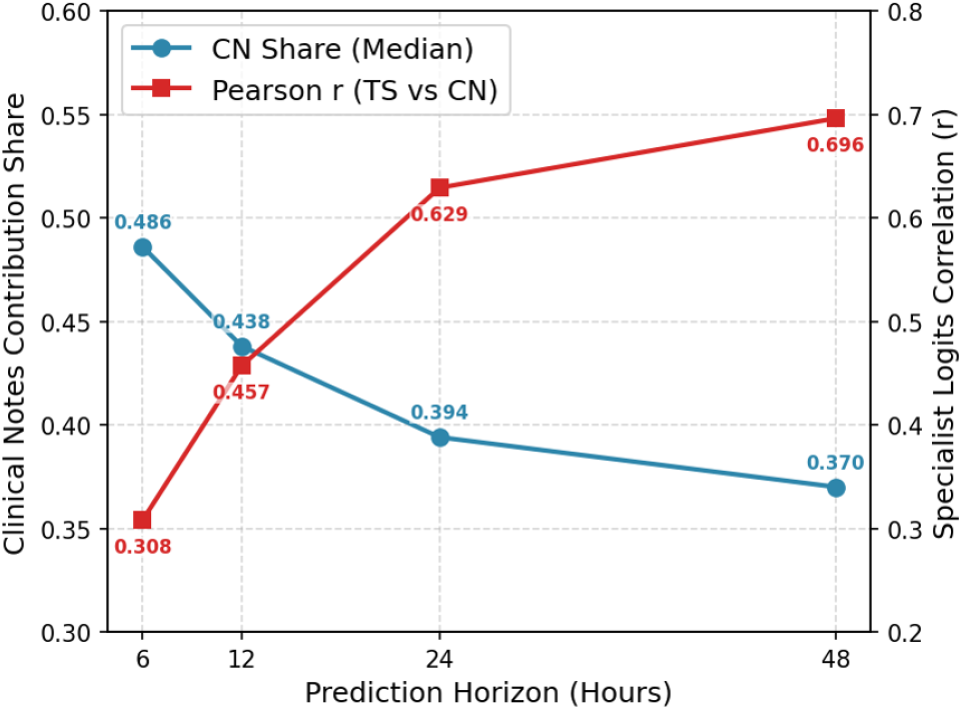
Modality complementarity across horizons. Median clinical-note contribution share (blue circles, left axis) increases as horizon shortens, from 0.370 at 48 h to 0.486 at 6 h, while Pearson correlation between specialist logits (red squares, right axis) decreases from 0.696 to 0.308. Together, these trends indicate increasing complementarity under temporal truncation: as physiological evidence becomes sparser, clinical notes contribute a larger relative share and the two specialists become less redundant.

**Figure 3:**
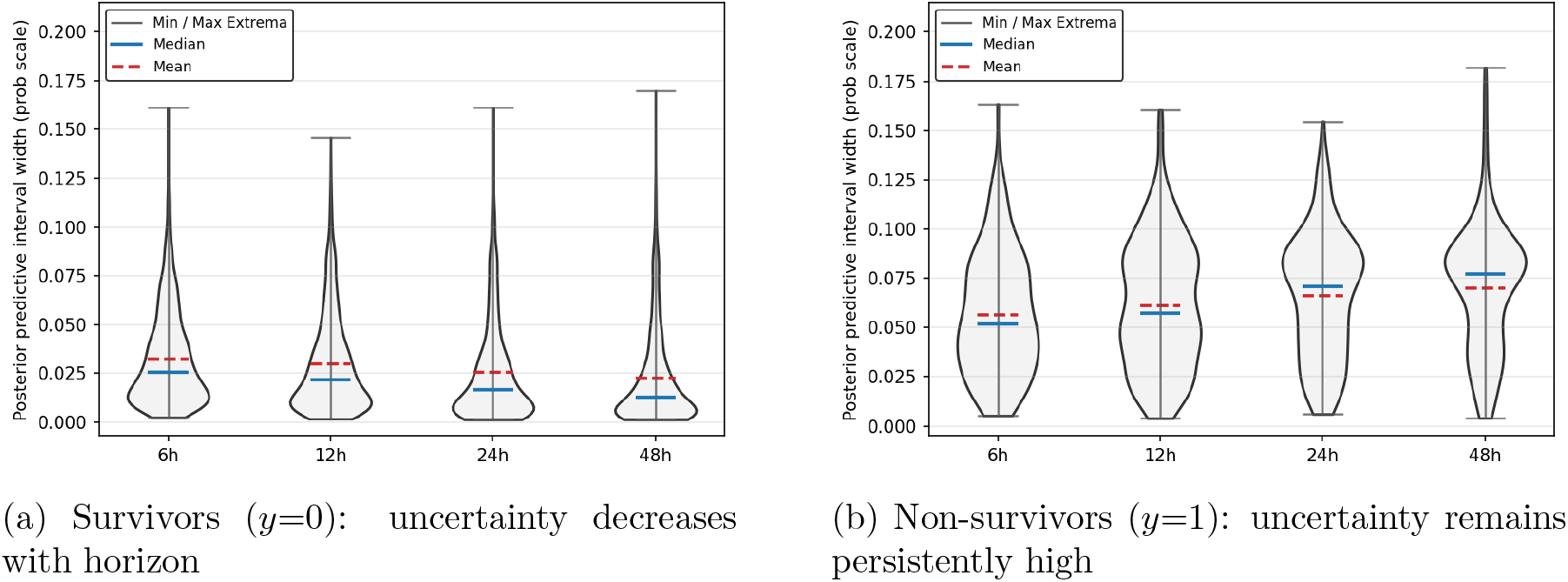
Class-stratified epistemic uncertainty by horizon (posterior predictive width q95–q5, probability scale). Survivors stabilize as physiological evidence accumulates; non-survivors do not. The class asymmetry becomes more pronounced at later horizons (median ratio 2.0 at 6h → 6.0 at 48h), flagging the cases the model finds hardest to classify: those whose fused specialist signals do not converge to a confident prediction regardless of observation window. Full per-class quantiles are in Appendix Table 6; overall horizon trend is in Appendix Figure 9.

The Pearson correlation between specialist logits falls from *r* = 0.696 at 48h to *r* = 0.308 at 6h (Figure 2), with variance explained dropping from 48.4% to 9.5%, indicating a transition from largely overlapping to largely distinct signals as fewer physiological hours are available. This declining correlation provides a model-grounded explanation for the preserved fusion margin observed in Section 3.1: decorrelated specialists produce more diverse error patterns, making their combination more effective Kuncheva and Whitaker (2003). Figure 2 shows the joint trend directly: CN share rises as specialist correlation falls, with the two curves crossing near 12h. Both modalities should therefore be retained for early-horizon evaluation; dropping either would incur the largest complementarity cost precisely when physiological data are scarcest. Episode-level dominance patterns are consistent with this aggregate trend: balanced cases (CN share 45–55%) increase from 11.5% at 48h to 19.0% at 6h, TS-dominant cases fall from 16.7% to 10.0% (Appendix Figure 8), and the proportion of both-high specialist consensus cases (Agree High: both predict *p >* 0.5) declines from 5.1% to 1.8% as fewer physiological hours are available to drive both specialists into high-mortality agreement (Appendix Table 3).

#### Signed contributions

Modality share and logit correlation describe aggregate complementarity; signed contributions reveal the directional mechanism: time-series pushes most patients toward low-risk with growing force as data accumulate, while clinical notes maintain a distinctive positive tail that drives high-risk identifications at all horizons.

To evaluate late-fusion behavior across horizons, we analyzed signed modality contributions from the logistic-regression stacker at 6, 12, 24, and 48 hours. Because coefficients were converted from standardized-logit space to effective raw-logit coefficients, fused predictions decompose exactly as:

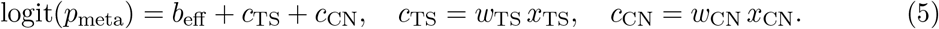

Appendix Figure 7 shows the patient-level contribution distributions at each horizon. With ≈11% in-hospital mortality, central tendency is below zero for both modalities throughout. For time-series, contributions grow more negative and more dispersed across all percentiles as physiology accumulates; the p95 remains near zero at all horizons, confirming the LSTM rarely contributes strong high-risk signal. For clinical notes, the positive tail strengthens: the fraction above zero rises from 10.2% at 6h to 19.8% at 48h.

TS is attenuated at early horizons, CN remains anchored throughout; this divergence is the per-episode manifestation of the complementarity pattern in Section 3.2. Full per-quantile statistics are in Appendix Table 4.

### 3.3. Per-Case Epistemic Uncertainty and Its Class Asymmetry

The preceding sections establish that the frozen LR-based fusion remains effective and interpretable across early horizons. We next ask a different question: how much confidence should be placed in any individual fused prediction? The NUTS extension of the LR stacker provides this: by treating fusion weights as random variables, it produces a posterior predictive distribution per episode whose width (*u*_*h*_, Eq. 2) quantifies how much the predicted probability varies under uncertainty about the fusion parameters.

#### NUTS recovers identical LR discrimination

The Bayesian (NUTS) meta-stacker yields essentially identical discrimination to the deterministic LR meta-stacker at every horizon (AUROC and AUPRC differences ≤ 0.001; full comparison in Appendix Table 5), confirming that uncertainty quantification is added without any performance trade-off. This is expected for a 3-parameter model, where the posterior predictive mean closely tracks the maximum-likelihood solution while the posterior over weights enables per-case interval estimation.

#### Overall uncertainty

Using Eq. (2) with a fixed posterior learned at 48 h, we quantified horizon-wise posterior predictive interval widths (q95–q5) on the probability scale. Median posterior predictive width increases from 0.0156 at 48 h to 0.0282 at 6 h (1.81×); corresponding ratios at 12h and 24h are 1.59× and 1.26× respectively (see Appendix Figure 9). Notably, the upper tail (q90–q95) remains comparatively stable across horizons, suggesting that a subset of episodes sustain high epistemic uncertainty regardless of available data, motivating the class-stratified analysis below.

#### Survivors stabilize and non-survivors remain uncertain

Label-stratified distributions reveal that the overall horizon trend is driven primarily by the negative class. For *y* = 0 (survivors), median uncertainty decreases as horizon increases. For *y* = 1 (non-survivors), uncertainty is consistently higher and its median increases with horizon, producing a class asymmetry that becomes more pronounced at later horizons.

Appendix Table 6 gives full quantiles by class and horizon. The ratio of median uncertainty for non-survivors to survivors increases from 2.02 at 6 h to 5.96 at 48 h. By the time maximum physiological evidence is available, the model is substantially more certain about survivors while its uncertainty about non-survivors has barely moved from its 6h level.

This asymmetry has a specific model-level reading. For survivors, additional ICU monitoring provides increasingly consistent physiological evidence that the specialist logits integrate into a confident low-risk prediction. For non-survivors, the fused specialist signals do not converge: the posterior remains wide because no stable configuration of the two weights produces a consistently high-risk output for these episodes; this is not a model failure, but an accurate signal of inherent classification difficulty. What the Bayesian stacker makes explicit, and the LR point estimate conceals, is that difficulty, per episode and per horizon.

#### Operability annotation

The practical question is whether high epistemic uncertainty merely reflects data scarcity, which would make it redundant with the point estimate, or whether it preferentially concentrates on episodes the model finds hardest to classify. Across all horizons, high-uncertainty episodes are substantially enriched for non-surviving patients. At 48 h, the top 10% most-uncertain episodes contain non-surviving patients at 4.90× the base rate; the pattern holds consistently from 6h (3.05×) through 48h. This confirms that epistemic uncertainty carries information beyond the point estimate: it concentrates on the cases where the model’s fused specialist signals fail to converge, rather than on cases that are simply data-sparse.

However, this also cautions against naive uncertainty-gated abstention in a highly imbalanced setting: deferring on high-uncertainty cases would disproportionately defer non-surviving patients. We therefore treat epistemic uncertainty as an operability annotation, a flag that a given prediction warrants closer scrutiny, rather than as a deployed deferral or triage policy.

### 3.4. Clinical Notes Persistence as Stable Contextual Inputs

We hypothesized that clinical notes encode stable admission-level context, including presenting diagnosis, clinician assessment, and comorbidities, that remains predictive between documentation events. If so, LOCF should suffice without staleness penalties. We tested this against two decay-assuming alternatives (Section 2.7).

#### 3.4.1. LOCF Baseline: Stepwise Gains at Documentation Events

Using the fixed 6h cohort (*N* =11,881) evaluated hourly from hour 6 through hour 48, the LOCF ensemble achieves AUROC 0.777 at 6h and rises to 0.910 at 48h; AUPRC rises from 0.306 to 0.584 over the same range (Figure 4). The trajectories display a characteristic step pattern: performance accelerates at documentation events (hours 12, 24, and 48, when a new cumulative note is incorporated), and accumulates gradually between events from time-series accumulation alone. Carrying a note forward across an inter-event interval does not harm prediction; the ensemble maintains or improves at every between-event hour, confirming the LOCF assumption: a note written at hour 6 remains informative at hour 11.

**Figure 4:**
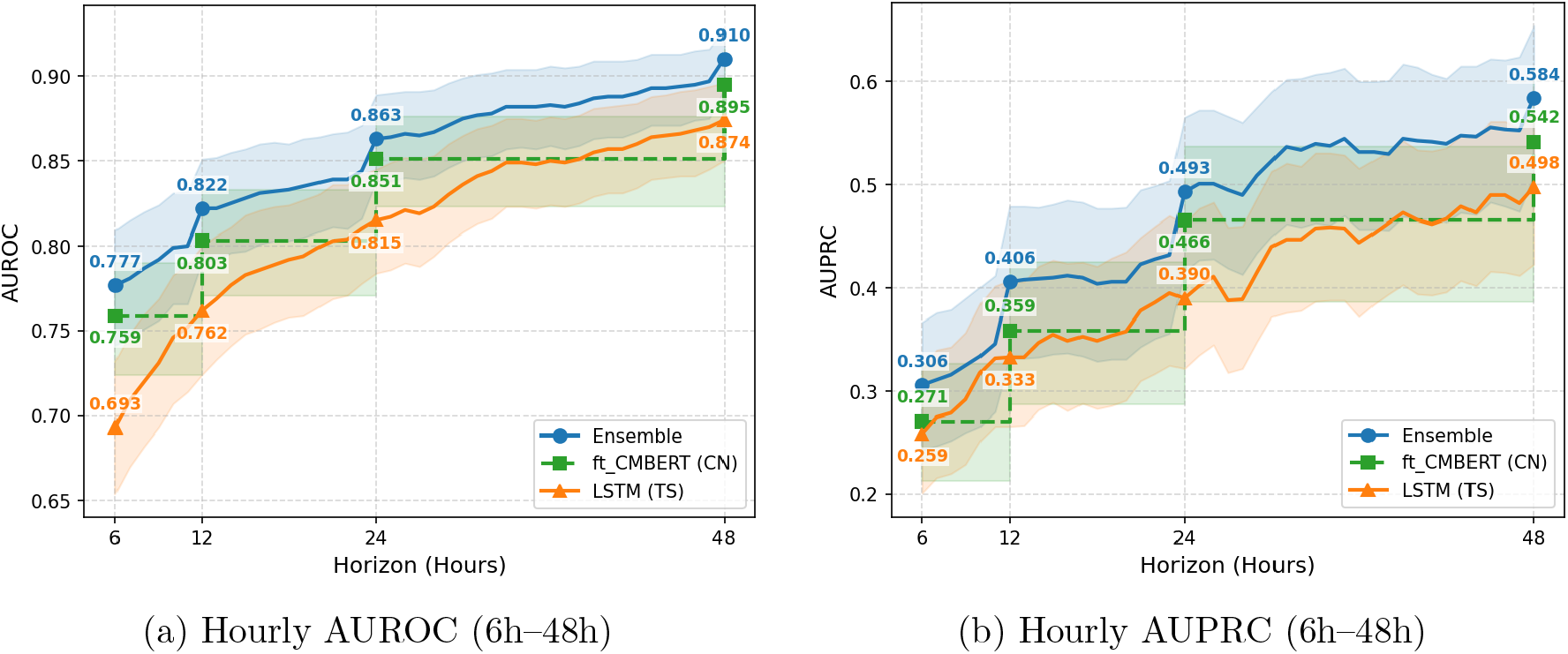
Continuous hourly discrimination under LOCF on the fixed 6h cohort (*N* =11,881). Shaded bands show 95% bootstrap confidence intervals. Ensemble (blue), ft-CMB specialist (green, dashed), LSTM specialist (orange). Performance rises at documentation events (hours 12, 24, and 48; vertical step increases) and accumulates gradually between events from physiological data alone. The ensemble consistently exceeds both specialists at every hour. Carrying the most recent note forward does not degrade prediction between documentation events, consistent with the note-persistence hypothesis.

##### Exponential decay

To test whether note contributions should diminish as time since documentation grows, we applied exponential decay to the CN logit at inference time: logit(*p*_meta_) = *b*_eff_ + *w*_TS_*x*_TS_ + *w*_CN_ · *e*^−*α*Δ*h*^ · *x*_CN_, where Δ*h* is hours since the most recent note. Three decay rates were tested (half-life 6h, 12h, 24h) as post-hoc rescaling without retraining. All three variants perform within or below the LOCF confidence band at all evaluation hours; stronger decay consistently reduces AUROC and AUPRC at hours distant from documentation events, precisely the intervals where the staleness hypothesis predicts the largest benefit.

##### Time-since-note

Documentation timing could still be informative independent of note content, as a proxy for monitoring intensity or care escalation patterns. To test this, we extended the stacker with Δ*h* as a third meta-feature and retrained on validation predictions: logit(*p*_meta_) = *b*_eff_ + *w*_TS_*x*_TS_ + *w*_CN_*x*_CN_ + *w*_Δ_Δ*h*. The learned coefficient *w*_Δ_ ≈ 0.01 is effectively zero, and performance is indistinguishable from the two-feature LOCF baseline. Across all three tests, LOCF is sufficient: clinical notes encode admission-level context that operates on a different informational timescale from hourly physiology, and prediction quality was not sensitive to documentation timing within the tested window.

## 4. Discussion

In this work, we examined whether a transparent multimodal ICU mortality ensemble trained once at 48 h can be used earlier without retraining, and whether its predictions remain trustworthy under that zero-shot horizon shift. The main finding is that early-horizon use does not reduce trustworthiness to discrimination alone: the LR-based ensemble retained both discrimination and strong calibration across 6–48 h while consistently outperforming both unimodal specialists. The calibration result is particularly important because it is empirical under horizon shift rather than guaranteed by design. The LR stacker was fitted on the 48h training distribution; applying the same frozen fusion rule at earlier horizons changes the specialist logits it receives, so calibration could have degraded substantially. Instead, ECE remained ≤ 0.038 at all horizons, whereas both specialists required post-hoc recalibration to approach comparable reliability. While calibration remains underreported in the multimodal ICU mortality prediction literature, this shows that probabilistic reliability can be preserved under zero-shot horizon transfer in a transparent late-fusion architecture.

The preserved fusion advantage at earlier horizons has a model-grounded explanation in changing modality complementarity. As horizon shortens, the time-series specialist degrades faster than the notes specialist, while the ensemble remains above both. This is accompanied by two consistent shifts in fusion behavior: the median clinical-notes contribution share rises from 0.370 at 48 h to 0.486 at 6 h, and the Pearson correlation between specialist logits falls from *r* = 0.696 to *r* = 0.308, reducing shared variance from 48.4% to 9.5%. The two specialists therefore become less redundant when physiological evidence is sparse; lower redundancy implies more diverse error structure and a greater fusion benefit Kuncheva and Whitaker (2003). The signed-contribution analysis confirms this at episode level: the time-series specialist (TS) increasingly resolves the low-risk majority, while the clinical notes specialist (CN) maintains a persistent positive tail, an attribution that is algebraically exact because the LR stacker is linear.

The Bayesian NUTS extension shows that population-level performance is an incomplete account of trustworthiness. What it contributes is per-case epistemic reliability. The key finding is a class-level asymmetry: survivors stabilize as evidence accumulates while non-survivors remain persistently uncertain, a pattern not explained by data sparsity, because it persists when input evidence is maximal. The model becomes progressively more certain about episodes whose fused signals support survival, while remaining uncertain about episodes whose specialist signals do not converge. Epistemic uncertainty therefore functions as an operability annotation, a per-case flag that a prediction warrants closer scrutiny, not a justification for uncertainty-gated abstention, which in an imbalanced setting would disproportionately defer non-surviving patients.

The continuous hourly analyses support the note-persistence hypothesis. All three staleness tests converge on the same conclusion: clinical notes encode admission-level context that operates on a different informational timescale from hourly physiology and is not superseded by vital sign evolution within the first 48 h. Within the tested window, prediction quality was not sensitive to documentation timing, which may simplify operational requirements for systems of this type.

### Limitations

First, evaluation is conducted on a single institution’s data (MIMIC-III); generalizability to other hospitals, electronic health record systems, and patient populations is unverified. Second, decision-curve analysis is not reported and net clinical benefit has not been characterized; these results should not be interpreted as evidence of clinical utility. Third, the zero-shot evaluation tests a frozen 48h-trained system; models retrained at each horizon may achieve higher performance, and that comparison is not performed here. Fourth, epistemic uncertainty is quantified only at the fusion layer; aleatoric uncertainty and specialist-level uncertainty are not modeled. Fifth, cohort construction requires at least one clinical note within the first *h* hours; settings with sparser early documentation may exhibit different note-persistence behavior.

## 5. Conclusion

This paper provides an answer to a practical translational question: whether a transparent multimodal ICU mortality model trained at 48 h can be meaningfully characterized at earlier horizons without retraining. Within that zero-shot setting, trustworthiness proved characterizable across four measurable dimensions: discrimination, calibration, fusion behavior, and per-case uncertainty, without any retraining.

Taken together, the frozen ensemble preserved calibration and discrimination across 6– 48 h; its fusion advantage was accompanied by increasing modality complementarity; its Bayesian extension revealed a persistent uncertainty asymmetry concentrated in hard non-survivor cases; and clinical notes behaved as a stable contextual input rather than a decaying signal. Rather than treating early prediction as an accuracy-only problem, this paper shows that early-horizon multimodal prediction can be characterized as a trustworthiness problem, with explicit evidence on when reliable signal emerges, how fusion behaves when physiology is sparse, and which cases remain intrinsically hard to classify.

## Data Availability

The data used in this study are available from the MIMIC-III clinical database (https://physionet.org/content/mimiciii/1.4/) subject to completion of required credentialing and data use agreements.

## Appendix A. Ensemble Discrimination Curves

**Figure 5:**
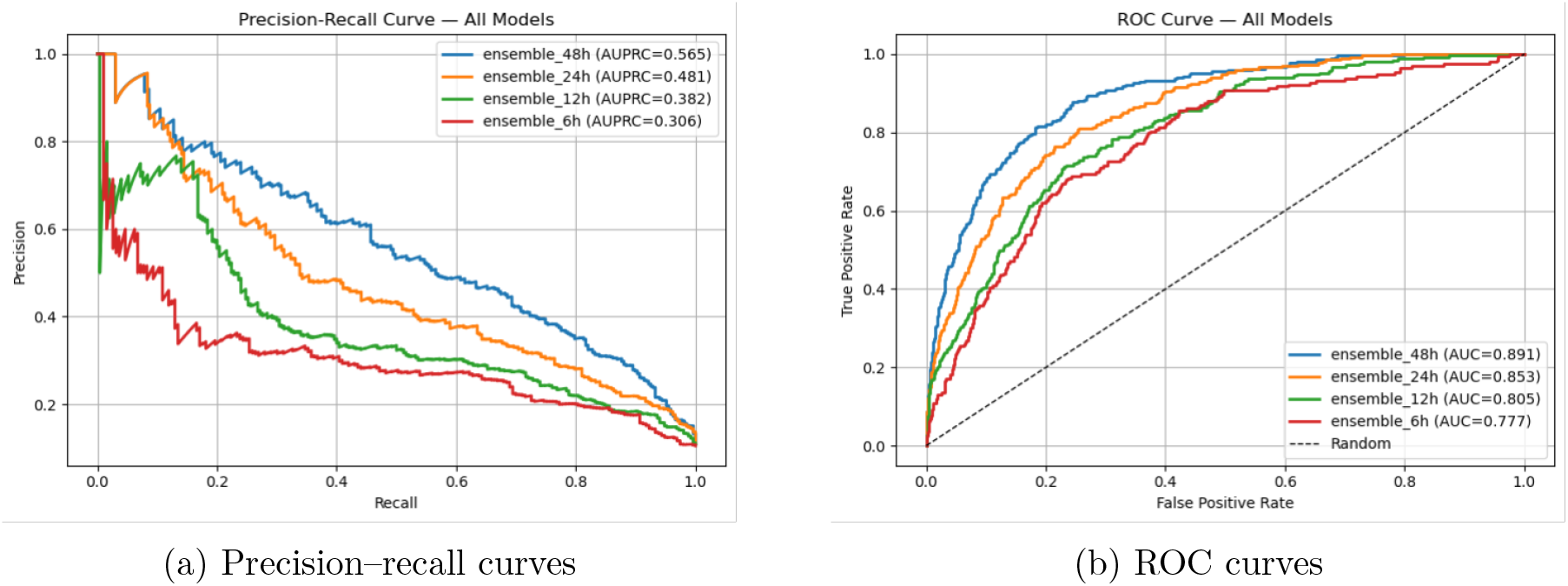
Precision–recall (left) and ROC (right) curves for the multimodal LR ensemble at 6, 12, 24, and 48 hours on the held-out test set. AUPRC and AUROC values match Table 2.

## Appendix B. Stacking Algorithm Comparison

**Figure 6:**
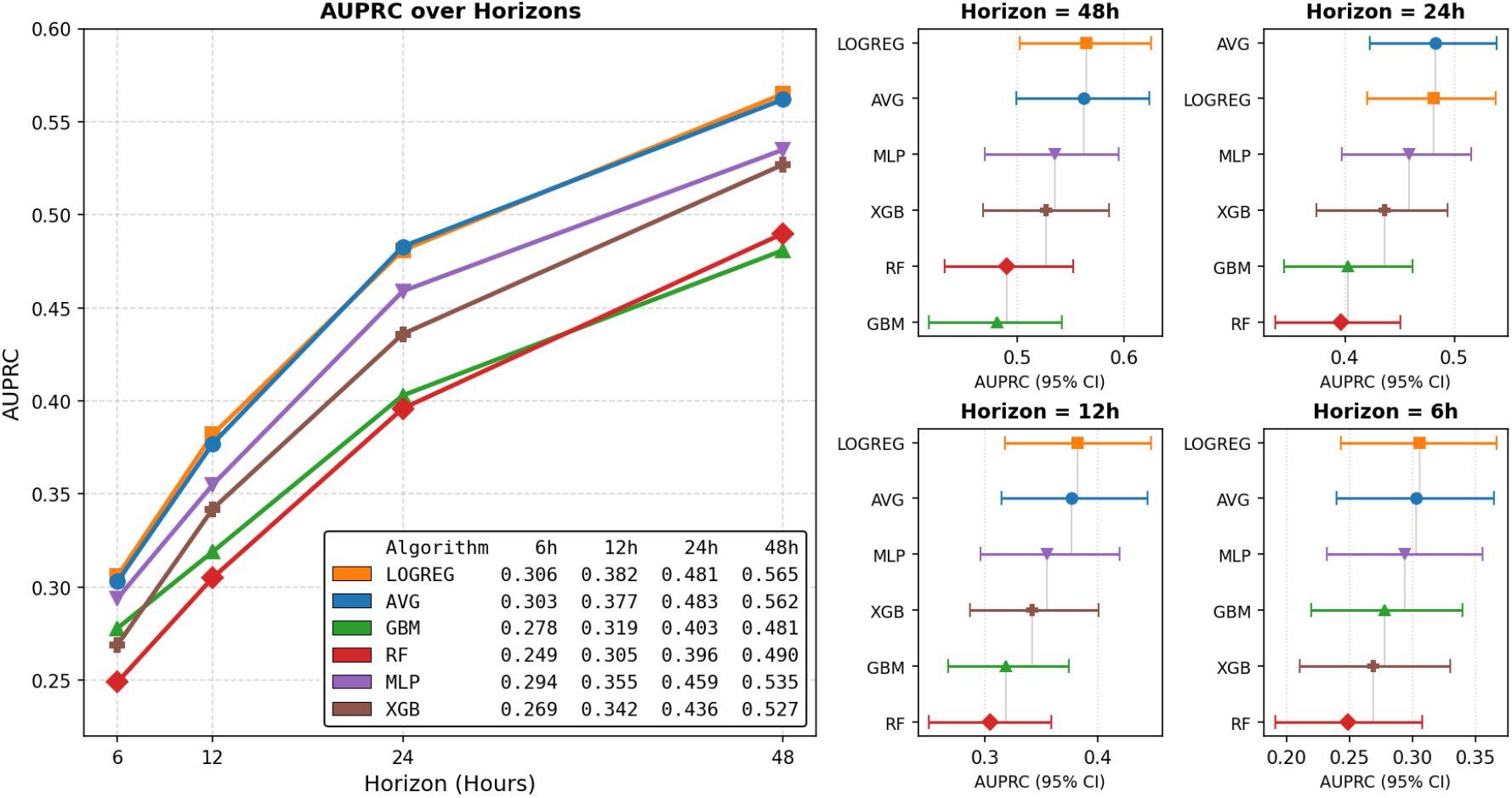
Stacking algorithm comparison on held-out test AUPRC. Left: AUPRC trajectories across horizons for six meta-learner candidates (LOGREG, AVG, GBM, RF, MLP, XGB). Right: per-horizon forest plots with 95% bootstrap confidence intervals. LOGREG and AVG consistently match or outperform all other candidates at every horizon; tree-based and neural alternatives show lower point estimates and wider intervals, consistent with variance induction from an underdetermined 2-feature input.

## Appendix C. Signed Modality Contribution Distributions

**Figure 7:**
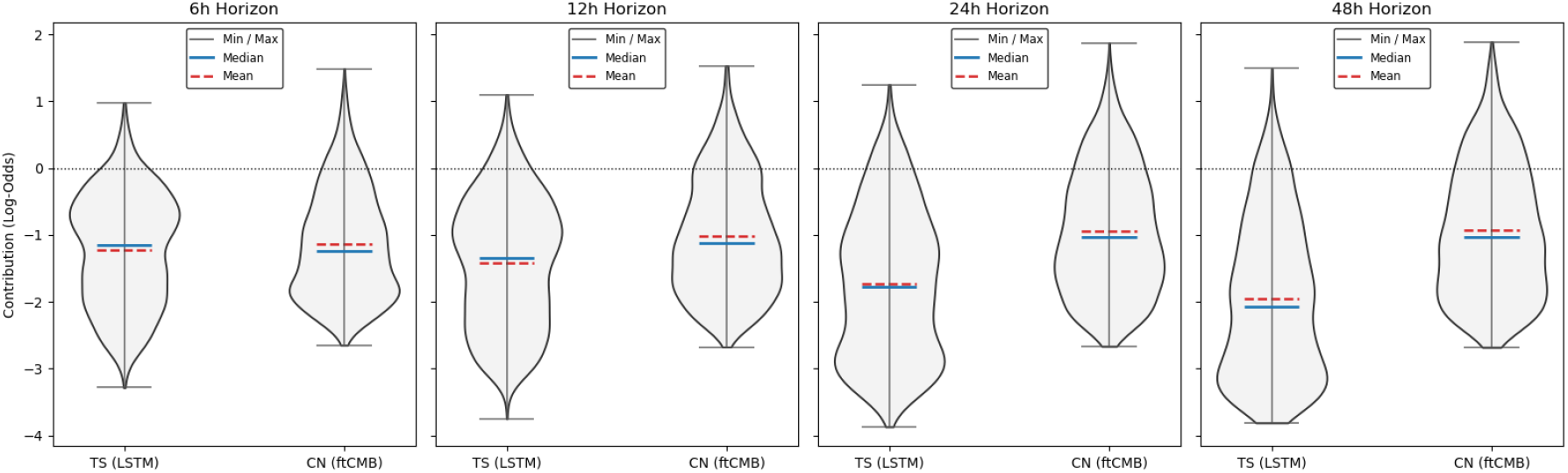
Signed modality contribution distributions across decision horizons (LR stacker). Each violin shows patient-level weighted contributions in raw log-odds units for the LSTM (TS) and ClinicalModernBERT (CN) specialists. The dotted line at zero marks no directional effect. Values below zero push the ensemble toward lower mortality odds; values above zero push toward higher mortality odds. Blue solid = median; red dashed = mean. TS contributions grow more negative and more dispersed with time; CN contributions maintain a distinctive positive tail at all horizons.

## Appendix D. Modality Dominance Patterns

**Figure 8:**
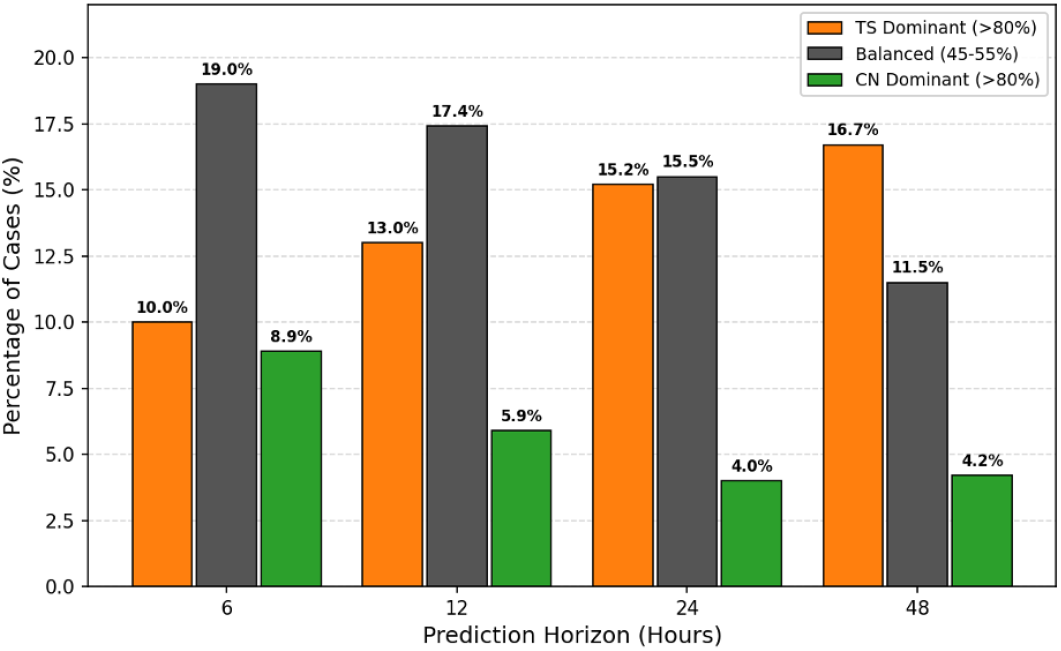
Episode-level modality dominance patterns by horizon. TS Dominant: CN share *<* 20%; Balanced: CN share 45–55%; CN Dominant: CN share *>* 80%. At earlier horizons, TS-dominant cases decrease (16.7%→10.0%) and balanced cases increase (11.5%→19.0%), consistent with increasing two-modality fusion at early horizons.

## Appendix E. Specialist Agreement Patterns

**Table 3:** Specialist agreement patterns across horizons (test set). Agree High: both specialists predict *p >* 0.5; Conflict: one above, one at or below 0.5; Agree Low: both predict *p* ≤ 0.5. Counts and row percentages shown.

| Horizon | Agree High ($\uparrow\uparrow$) | Conflict ($\uparrow\downarrow$) | Agree Low ($\downarrow\downarrow$) |
| --- | --- | --- | --- |
| 48h | 124 (5.1%) | 394 (16.1%) | 1930 (78.8%) |
| 24h | 105 (4.4%) | 399 (16.7%) | 1892 (79.0%) |
| 12h | 72 (3.1%) | 346 (15.1%) | 1874 (81.8%) |
| 6h | 33 (1.8%) | 213 (11.6%) | 1591 (86.6%) |

## Appendix F. Signed Modality Contribution Statistics

**Table 4:** Summary statistics for signed modality contribution distributions (LR stacker). All values in raw log-odds units. Positive fraction: proportion of episodes where the modality contributes a positive (mortality-increasing) log-odds term.

| Horizon | Modality | Mean | Median | SD | p5 | p25 | p75 | p95 | Pos. fraction |
| --- | --- | --- | --- | --- | --- | --- | --- | --- | --- |
| 6h | TS (LSTM) | -1.220 | -1.153 | 0.830 | -2.570 | -1.899 | -0.578 | -0.002 | 5.0% |
|  | CN (ft-CMB) | -1.132 | -1.243 | 0.804 | -2.233 | -1.809 | -0.568 | 0.295 | 10.2% |
| 12h | TS (LSTM) | -1.413 | -1.345 | 0.952 | -2.940 | -2.208 | -0.678 | 0.114 | 6.3% |
|  | CN (ft-CMB) | -1.013 | -1.121 | 0.866 | -2.250 | -1.712 | -0.367 | 0.493 | 15.1% |
| 24h | TS (LSTM) | -1.731 | -1.780 | 1.097 | -3.311 | -2.714 | -0.896 | 0.154 | 6.9% |
|  | CN (ft-CMB) | -0.940 | -1.038 | 0.926 | -2.282 | -1.685 | -0.240 | 0.636 | 18.5% |
| 48h | TS (LSTM) | -1.960 | -2.069 | 1.160 | -3.518 | -2.994 | -1.085 | 0.101 | 6.5% |
|  | CN (ft-CMB) | -0.936 | -1.038 | 0.975 | -2.314 | -1.754 | -0.194 | 0.720 | 19.8% |

## Appendix G. NUTS vs. LR Discrimination Parity

**Table 5:** Discrimination performance: Bayesian NUTS vs. deterministic LR meta-stacker. Differences are ≤0.001 at all horizons, confirming the Bayesian extension is non-destructive.

| Metric | NUTS | | LR | | $\Delta$ |
| --- | --- | --- | --- | --- | --- |
| *6h horizon* |  |  |  |  |  |
| AUROC | 0.776 | [0.741,0.809] | 0.777 | [0.741,0.809] | −0.001 |
| AUPRC | 0.306 | [0.243,0.367] | 0.306 | [0.243,0.367] | 0.000 |
| *12h horizon* |  |  |  |  |  |
| AUROC | 0.805 | [0.778,0.830] | 0.805 | [0.778,0.831] | 0.000 |
| AUPRC | 0.382 | [0.317,0.448] | 0.382 | [0.318,0.447] | 0.000 |
| *24h horizon* |  |  |  |  |  |
| AUROC | 0.853 | [0.831,0.875] | 0.853 | [0.831,0.875] | 0.000 |
| AUPRC | 0.481 | [0.420,0.538] | 0.481 | [0.420,0.538] | 0.000 |
| *48h horizon* |  |  |  |  |  |
| AUROC | 0.891 | [0.872,0.909] | 0.891 | [0.872,0.909] | 0.000 |
| AUPRC | 0.564 | [0.503,0.624] | 0.565 | [0.503,0.625] | −0.001 |

## Appendix H. Overall Epistemic Uncertainty by Horizon

**Figure 9:**
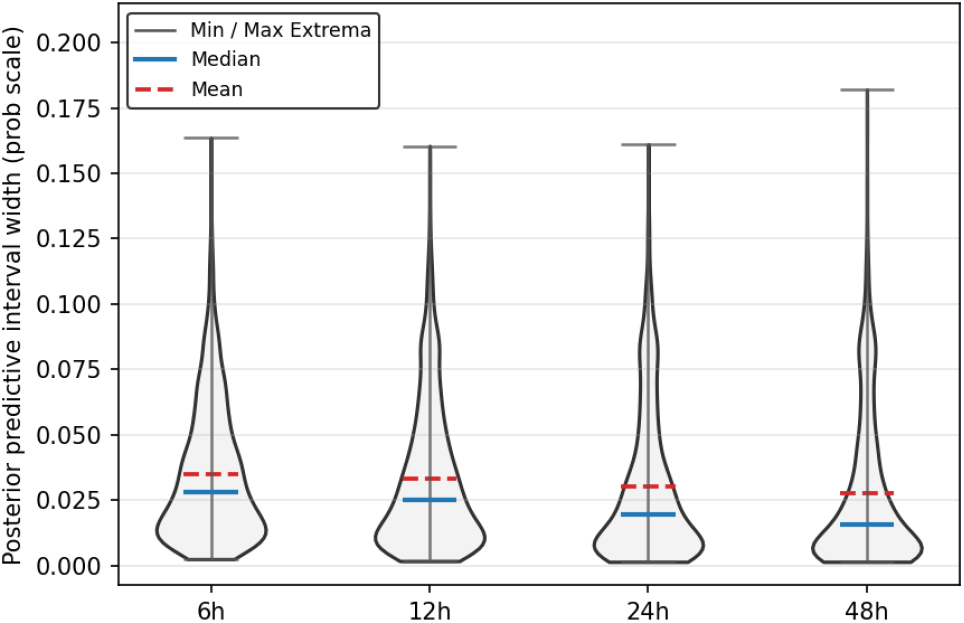
Epistemic uncertainty by horizon (all episodes). Posterior predictive interval width (q95–q5) on the probability scale, under a fixed posterior learned at 48 h. Median width increases 1.81× at 6h relative to 48h; the upper tail (q90–q95) remains stable, indicating a subset of persistently uncertain episodes characterized by the class-stratified analysis in Figure 3.

## Appendix I. Epistemic Uncertainty by Class and Horizon

**Table 6:** Posterior predictive interval width (q95–q5) by horizon and outcome class.

| Class | Horizon | Mean | Median | q90 | q95 |
| --- | --- | --- | --- | --- | --- |
| $y=0$ | 6h | 0.032567 | 0.025689 | 0.068168 | 0.079425 |
| $y=0$ | 12h | 0.029884 | 0.021966 | 0.068452 | 0.083485 |
| $y=0$ | 24h | 0.025799 | 0.016441 | 0.065811 | 0.083452 |
| $y=0$ | 48h | 0.022511 | 0.012959 | 0.060253 | 0.078685 |
| $y=1$ | 6h | 0.056440 | 0.051876 | 0.097837 | 0.111306 |
| $y=1$ | 12h | 0.061233 | 0.057353 | 0.103024 | 0.112199 |
| $y=1$ | 24h | 0.066291 | 0.071038 | 0.103363 | 0.120587 |
| $y=1$ | 48h | 0.070139 | 0.077181 | 0.101046 | 0.114001 |

## Appendix J. Calibration by Specialist Agreement Category

**Table 7:** Ensemble calibration stratified by specialist agreement category and horizon. Actual Rate: observed mortality rate. Predicted Prob: mean ensemble predicted probability. Calibration Error: absolute difference. The 6h Agree High subgroup (*n* = 33) shows the largest calibration error (0.231), consistent with sample size instability.

| Horizon | Category | $N$ | Actual Rate | Predicted Prob | Calib. Error |
| --- | --- | --- | --- | --- | --- |
| 48h | Agree High | 124 | 0.669 | 0.674 | 0.005 |
|  | Conflict | 394 | 0.279 | 0.311 | 0.032 |
|  | Agree Low | 1930 | 0.038 | 0.044 | 0.006 |
| 24h | Agree High | 105 | 0.543 | 0.648 | 0.105 |
|  | Conflict | 399 | 0.306 | 0.324 | 0.018 |
|  | Agree Low | 1892 | 0.044 | 0.052 | 0.008 |
| 12h | Agree High | 72 | 0.528 | 0.620 | 0.092 |
|  | Conflict | 346 | 0.249 | 0.331 | 0.082 |
|  | Agree Low | 1874 | 0.067 | 0.065 | 0.002 |
| 6h | Agree High | 33 | 0.394 | 0.625 | 0.231 |
|  | Conflict | 213 | 0.272 | 0.326 | 0.054 |
|  | Agree Low | 1591 | 0.077 | 0.072 | 0.005 |

